# Inflammatory profiles are associated with long COVID up to 6 months after illness onset: a prospective cohort study of individuals with mild to critical COVID-19

**DOI:** 10.1101/2023.06.14.23291395

**Authors:** Elke Wynberg, Alvin X. Han, Hugo D.G. van Willigen, Anouk Verveen, Lisa van Pul, Irma Maurer, Ester M. van Leeuwen, Joost G. van den Aardweg, Menno D. de Jong, Pythia Nieuwkerk, Maria Prins, Neeltje A. Kootstra, Godelieve J. de Bree, the RECoVERED Study Group

## Abstract

**Background:** After initial COVID-19 disease, immune dysregulation may persist and drive post-acute sequelae of COVID-19 (PASC). We described longitudinal trajectories of cytokines in adults up to 6 months following SARS-CoV-2 infection and explored early predictors of PASC.

**Methods:** RECoVERED is a prospective cohort of individuals with laboratory-confirmed SARS-CoV-2 infection between May 2020 and June 2021 in Amsterdam, the Netherlands. Serum was collected at weeks 4, 12 and 24 of follow-up. Monthly symptom questionnaires were completed from month 2 after illness onset onwards; lung diffusion capacity (DLCO) was tested at 6 months. Cytokine concentrations were analysed by human magnetic Luminex screening assay. We used a linear mixed-effects model to study log-concentrations of cytokines over time, assessing their association with socio-demographic and clinical characteristics that were included in the model as fixed effects.

**Results:** 186/349 (53%) participants had ≥2 serum samples and were included. Of these, 101 (54%: 45/101[45%] female, median age 55 years [IQR=45-64]) reported PASC at 12 and 24 weeks after illness onset. We included 37 reference samples (17/37[46%] female, median age 49 years [IQR=40-56]). PASC was associated with raised CRP and abnormal diffusion capacity with raised IL10, IL17, IL6, IP10 and TNFα at 24 weeks in the multivariate model. Early (0-4 week) IL-1**β** and BMI at illness onset were predictive of PASC at 24 weeks.

**Conclusions:** Our findings indicate that immune dysregulation plays an important role in PASC pathogenesis, especially among those individuals with reduced pulmonary function. Early IL-1**β** shows promise as predictors of PASC.

## Introduction

Almost half of individuals[1] infected with severe acute respiratory syndrome coronavirus 2 (SARS-CoV-2) are estimated to experience symptoms related to “long COVID” or post-acute sequelae of COVID-19 (PASC). There are many uncertainties regarding the pathophysiological processes and predictors of PASC. Numerous studies have suggested that hyperinflammation observed during acute infection among individuals with severe COVID-19 [2-4] may persist among those with ongoing symptoms[5]. However, less is known about the role of aberrant cytokine levels in the pathogenesis of PASC in individuals with initially mild or moderate COVID-19.

Observations to date have been largely inconclusive due to heterogeneity in research objectives, study design and population. Some studies aimed to outline immune disturbances in the first months after infection[6-8] in order to better understand COVID-19 immunopathology, whilst others specifically aimed to identify early inflammatory predictors for PASC that could be of clinical relevance [9-13]. In a prospective cohort of individuals with COVID-19, of whom 22% had been hospitalised, IL6 remained significantly raised among participants with PASC compared to those without PASC 90 days after infection[6]. A second study also observed elevated levels of IL6, as well as IL1 and TNF, around 10 months post symptom onset[14]. In a cohort of largely non-hospitalised patients, elevated TNFα at 4 and 8 months were strongest predictors of PASC overall, with no difference observed in IL6 between those with and without PASC[11]. Prospective data that aims to both outline longitudinal immune profiles among individuals with mild to critical COVID-19, and identify possible early markers of PASC, is needed to better understand previous findings.

Using data from the RECoVERED cohort study in Amsterdam, the Netherlands, we aim to gain insight into longitudinal immune profiles among individuals with COVID-19 and PASC. A substantial proportion of study participants reported at least one persistent COVID-19 symptom 3 months or longer after initial illness onset[15]. In the current study, we evaluate the impact of SARS-CoV-2 infection on cytokine levels up to 6 months after illness onset, assess the association of PASC with specific inflammatory profiles, and evaluate possible early determinants of PASC.

## Methods

### Study design and participant enrolment

RECoVERED is a prospective cohort study of adults aged 16-85 with SARS-CoV-2 infection between May 2020 and June 2021 in Amsterdam, the Netherlands. Full details of study procedures have been published elsewhere[1]. Briefly, participants were enrolled within 7 days of diagnosis (at the Public Health Service of Amsterdam) or hospital admission (at the Amsterdam University Medical Centres [UMC]). All participants had laboratory-confirmed SARS-CoV-2 infection. RECoVERED was approved by the medical ethical review board of the Amsterdam University Medical Centres (METC NL73759.018.20). All participants provided written informed consent.

For the current analysis, we included participants with samples from at least two different time-points and with follow-up of at least 3 months following SARS-CoV-2 infection. We additionally included reference u samples from SARS-CoV-2-uninfected, healthy individuals (i.e., no comorbidities) collected between March 2020 and November 2021. These samples were randomly selected from a prospective serologic surveillance cohort study among hospital healthcare workers in the Amsterdam UMC (S3 study; METC NL73478.029.20)[16].

### Surveys on socio-demographic, clinical and symptom-related data

During the first month of follow-up, trained study staff interviewed participants on the presence of 20 different COVID-19 symptoms, took physical measurements, and recorded participants’ past medical and socio-demographic characteristics. Recorded symptoms included: fatigue, cough, fever, rhinorrhoea, sore throat, dyspnoea, loss of smell and/or taste, chest pain, headache, abdominal pain, confusion, arthralgia, myalgia, loss of appetite, wheeze, skin rash, nausea and/or vomiting, diarrhoea, earache, and spontaneous bleeding. Between months 3 to 12 of follow-up, monthly online questionnaires on the presence of the same 20 COVID-19 symptoms were completed by participants. Lung function tests including diffusion capacity (D_LCO_) were performed at 1, 6 and 12 months after illness onset (detailed methods described elsewhere[17]).

### Biological sampling

Serum samples were collected at day 0 and 7 and subsequently at months 1, 3, 6, 9, 12, 18 and 24 of follow-up. Additional serum samples were collected per protocol within 24 hours of receiving a COVID-19 vaccination and 7 and 28 days following each COVID-19 vaccination. All serum samples were processed within 24 hours and stored at -80LC. In the present study, we defined three time-frames of sample collection for longitudinal analyses: 0-4, 9-12 and 21-24 weeks after illness onset. Post-vaccination samples were defined as those collected within 28 days after administration of a COVID-19 vaccine.

### Definitions

Illness onset was defined as the earliest day of COVID-19 symptoms were experienced for symptomatic patients, or date of SARS-CoV-2 diagnosis for asymptomatic patients. PASC was defined as reporting at least one COVID-19 symptom that, from illness onset, occurred within one month and continued beyond 12 weeks[3]; symptoms arising after one month from illness onset were not attributed to PASC. COVID-19 clinical severity was categorised according to WHO criteria [14]. BMI was coded in kg/m^2^ as: <25, underweight or normal weight; 25-29, overweight; ≥30, obese. Diffusion capacity (D_LCO_) at 6 months after illness onset was defined as impaired according to American Thoracic Society (ATS) European Respiratory Society guidelines[18], as described previously described[17].

### Inflammation marker assays

C-reactive protein (CRP), CD14, CD163, tumor necrosis factor (TNF)-α, interferon-γ-inducible protein 10 (IP-10)/CXCL10, monocyte chemoattractant protein (MCP)1/CCL2, interleukin (IL)1β IL2, IL6, IL10, IL13 and IL17A, concentrations were analyzed in serum by human magnetic luminex screening assay (LXSAHM-02 and LXSAHM-10; R&D Systems). Assays were performed according to the manufacturer’s instructions. Using immunofluorescence, titers of total antinuclear antibodies (ANA) as indicators of possible auto-immunity were determined for samples collected ≤4 weeks after illness onset.

### Statistical analysis

Socio-demographic, clinical and study characteristics were compared between included participants with and without PASC at 12 weeks after illness onset. To assess selection bias, we compared features of included and excluded participants. We used the Fisher’s exact test to evaluate the association between PASC and impaired diffusion capacity at 6 months after illness onset.

We presented box-and-whisker plots of median (IQR) log-concentrations of cytokines at 9-12 and 21-24 weeks after illness onset, comparing in univariable analyses: i) with reference samples from uninfected, healthy individuals and ii) between study participants with and without PASC at 12 and 24 weeks after illness onset. Median (IQR) log-concentrations were compared using the Mann-Whitney U-test with correction for multiple testing using the Bonferroni method. Comparisons were first performed using data from the whole cohort (mild to critical COVID-19) and subsequently restricted to participants with initially mild or moderate COVID-19. In order to additionally assess differences in cytokine levels using an objective marker of PASC, we compared median (IQR) log-concentrations of cytokines measured at 21-24 weeks between participants with and without an impaired diffusion capacity (D_LCO_) at 6 months after illness onset.

To assess factors cross-sectionally associated with cytokine levels after illness onset, we applied two linear mixed-effects models: the first at 3 months (serum collected at 9-12 weeks) and the second at 6 months (serum collected at 21-24 weeks) after illness onset. We modelled log concentrations of cytokines with time since illness onset as a random effect and as fixed effects: sex, age and clinical characteristics (i.e. body mass index [BMI], comorbidities – defined at illness onset), PASC status (at 12 and 24 weeks after illness onset in each model, respectively), and recent [<4 weeks] vaccination). In order to incorporate an objective measure of abnormal pathology following COVID-19, we substituted PASC status (based on self-reported symptoms) for impaired diffusion capacity (D_LCO_) in an additional model at 6 months after illness onset. Condition indices were computed to ensure that there was no collinearity among the variables (i.e., condition index<10). A correlation matrix of cytokines at 0-4, 9-12 and 21-24 weeks were used to help interpret the complexity of inter-marker associations.

We then performed random forest regression, a model-free machine-learning approach, to identify the early predictors of: (1) PASC at 24 weeks and (2) higher levels, at 21-24 weeks after illness onset, of CRP and IL-6. We chose to investigate predictors of raised pro-inflammatory cytokines at 6 months in order to include more objective outcome measures than the current PASC definition. We performed k-fold cross validation (k=5) to tune the hyperparameters of each random forest regressor. We used F1 scores and mean squared error as scoring functions of the random forest regressors used in (1) and (2)/(3) respectively. We then computed Shapley additive explanation values as measures of importance for the different predictors[19]. CRP and IL6 at 0-4 weeks were not individually included as predictors of their measurements at 21-24 weeks.

All statistical analyses were performed in Python using the statsmodels package (v. 0.13.2)[20], whilst the random forest regression analyses were performed in Python using the scikit-learn package (v. 1.1.3).

## Results

### Study population, PASC status and lung function (diffusion capacity)

Of 349 RECoVERED participants, 186 (53.3%) had at least two serum sampling moments and were included in the present study. Included participants were more likely to be male and have initially mild COVID-19 (Supplementary Table S1). 101/186 (54%; 45/101[45%] female, median age 55 years [IQR=45-64]) included participants reported PASC at 12 weeks after illness onset (Table 1), of whom none recovered or were lost to follow-up between weeks 12 and 24. A subgroup (72/186; 38.7%) of participants had a diffusion capacity at 6 months after illness onset. Among the 22/72 who exhibited impaired diffusion capacity, more than half (12/22; 54.5%) also reported PASC at 6 months (p=0.031).

**Table 1.**
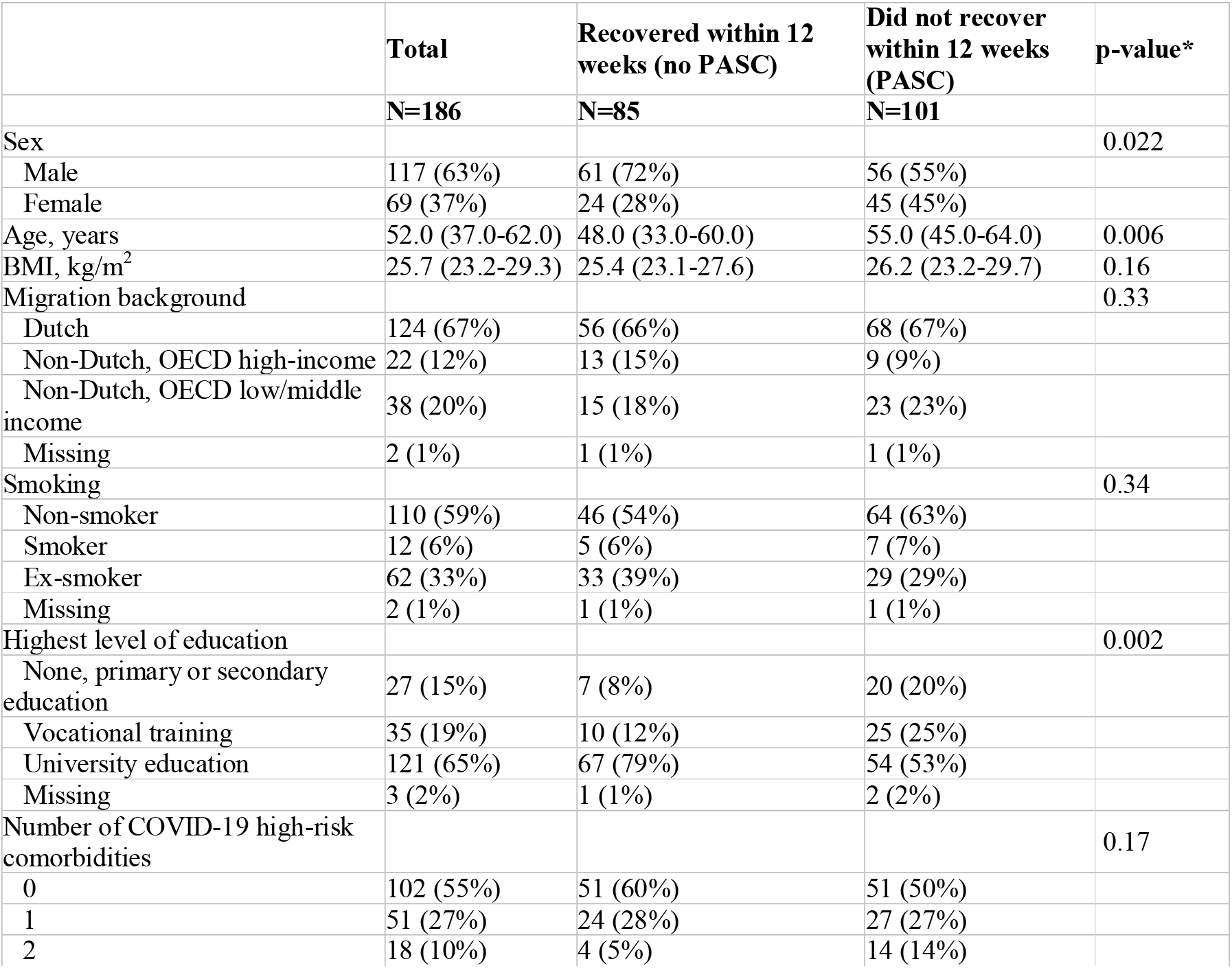
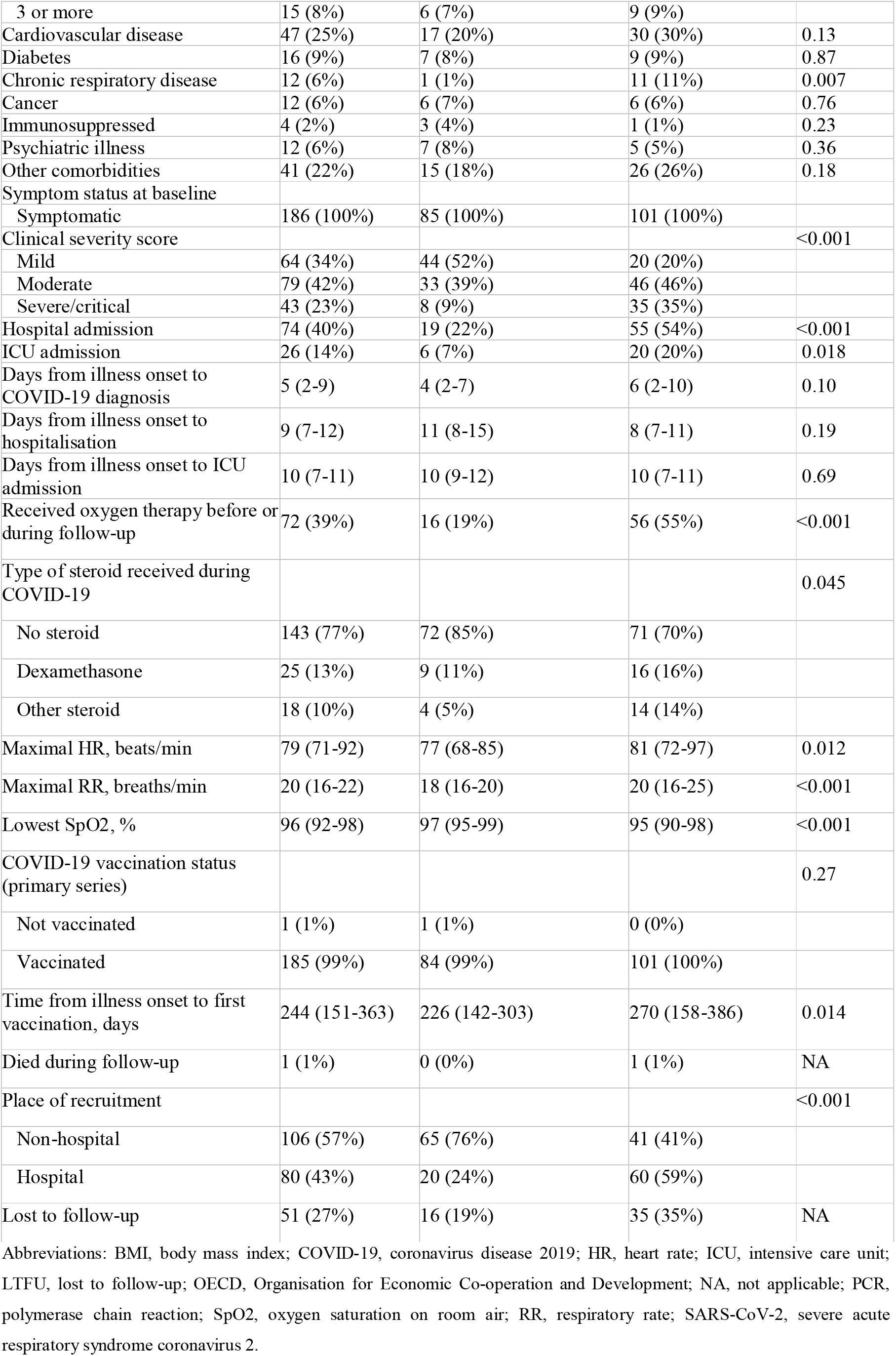
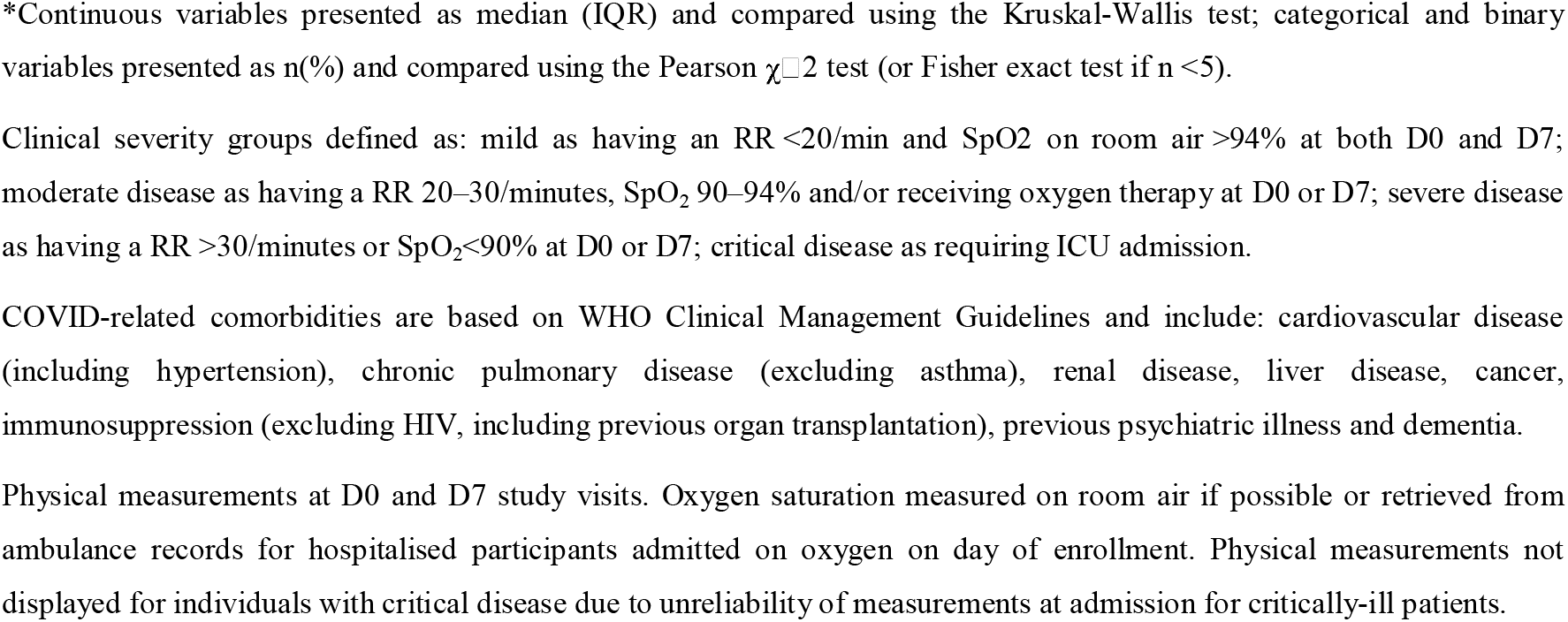
Socio-demographic, clinical and COVID-19-related characteristics of RECoVERED participants included in the current analyses by PASC status at 3 months after illness onset.

|  | Total | Recovered within 12 weeks (no PASC) | Did not recover within 12 weeks (PASC) | p-value* |
| --- | --- | --- | --- | --- |
|  | N=186 | N=85 | N=101 |  |
| Sex |  |  |  | 0.022 |
| Male | 117 (63%) | 61 (72%) | 56 (55%) |  |
| Female | 69 (37%) | 24 (28%) | 45 (45%) |  |
| Age, years | 52.0 (37.0-62.0) | 48.0 (33.0-60.0) | 55.0 (45.0-64.0) | 0.006 |
| BMI, kg/m<sup>2</sup> | 25.7 (23.2-29.3) | 25.4 (23.1-27.6) | 26.2 (23.2-29.7) | 0.16 |
| Migration background |  |  |  | 0.33 |
| Dutch | 124 (67%) | 56 (66%) | 68 (67%) |  |
| Non-Dutch, OECD high-income | 22 (12%) | 13 (15%) | 9 (9%) |  |
| Non-Dutch, OECD low/middle income | 38 (20%) | 15 (18%) | 23 (23%) |  |
| Missing | 2 (1%) | 1 (1%) | 1 (1%) |  |
| Smoking |  |  |  | 0.34 |
| Non-smoker | 110 (59%) | 46 (54%) | 64 (63%) |  |
| Smoker | 12 (6%) | 5 (6%) | 7 (7%) |  |
| Ex-smoker | 62 (33%) | 33 (39%) | 29 (29%) |  |
| Missing | 2 (1%) | 1 (1%) | 1 (1%) |  |
| Highest level of education |  |  |  | 0.002 |
| None, primary or secondary education | 27 (15%) | 7 (8%) | 20 (20%) |  |
| Vocational training | 35 (19%) | 10 (12%) | 25 (25%) |  |
| University education | 121 (65%) | 67 (79%) | 54 (53%) |  |
| Missing | 3 (2%) | 1 (1%) | 2 (2%) |  |
| Number of COVID-19 high-risk comorbidities |  |  |  | 0.17 |
| 0 | 102 (55%) | 51 (60%) | 51 (50%) |  |
| 1 | 51 (27%) | 24 (28%) | 27 (27%) |  |
| 2 | 18 (10%) | 4 (5%) | 14 (14%) |  |
| 3 or more | 15 (8%) | 6 (7%) | 9 (9%) |  |
| Cardiovascular disease | 47 (25%) | 17 (20%) | 30 (30%) | 0.13 |
| Diabetes | 16 (9%) | 7 (8%) | 9 (9%) | 0.87 |
| Chronic respiratory disease | 12 (6%) | 1 (1%) | 11 (11%) | 0.007 |
| Cancer | 12 (6%) | 6 (7%) | 6 (6%) | 0.76 |
| Immunosuppressed | 4 (2%) | 3 (4%) | 1 (1%) | 0.23 |
| Psychiatric illness | 12 (6%) | 7 (8%) | 5 (5%) | 0.36 |
| Other comorbidities | 41 (22%) | 15 (18%) | 26 (26%) | 0.18 |
| Symptom status at baseline |  |  |  |  |
| Symptomatic | 186 (100%) | 85 (100%) | 101 (100%) |  |
| Clinical severity score |  |  |  | <0.001 |
| Mild | 64 (34%) | 44 (52%) | 20 (20%) |  |
| Moderate | 79 (42%) | 33 (39%) | 46 (46%) |  |
| Severe/critical | 43 (23%) | 8 (9%) | 35 (35%) |  |
| Hospital admission | 74 (40%) | 19 (22%) | 55 (54%) | <0.001 |
| ICU admission | 26 (14%) | 6 (7%) | 20 (20%) | 0.018 |
| Days from illness onset to COVID-19 diagnosis | 5 (2-9) | 4 (2-7) | 6 (2-10) | 0.10 |
| Days from illness onset to hospitalisation | 9 (7-12) | 11 (8-15) | 8 (7-11) | 0.19 |
| Days from illness onset to ICU admission | 10 (7-11) | 10 (9-12) | 10 (7-11) | 0.69 |
| Received oxygen therapy before or during follow-up | 72 (39%) | 16 (19%) | 56 (55%) | <0.001 |
| Type of steroid received during COVID-19 |  |  |  | 0.045 |
| No steroid | 143 (77%) | 72 (85%) | 71 (70%) |  |
| Dexamethasone | 25 (13%) | 9 (11%) | 16 (16%) |  |
| Other steroid | 18 (10%) | 4 (5%) | 14 (14%) |  |
| Maximal HR, beats/min | 79 (71-92) | 77 (68-85) | 81 (72-97) | 0.012 |
| Maximal RR, breaths/min | 20 (16-22) | 18 (16-20) | 20 (16-25) | <0.001 |
| Lowest SpO2, % | 96 (92-98) | 97 (95-99) | 95 (90-98) | <0.001 |
| COVID-19 vaccination status (primary series) |  |  |  | 0.27 |
| Not vaccinated | 1 (1%) | 1 (1%) | 0 (0%) |  |
| Vaccinated | 185 (99%) | 84 (99%) | 101 (100%) |  |
| Time from illness onset to first vaccination, days | 244 (151-363) | 226 (142-303) | 270 (158-386) | 0.014 |
| Died during follow-up | 1 (1%) | 0 (0%) | 1 (1%) | NA |
| Place of recruitment |  |  |  | <0.001 |
| Non-hospital | 106 (57%) | 65 (76%) | 41 (41%) |  |
| Hospital | 80 (43%) | 20 (24%) | 60 (59%) |  |
| Lost to follow-up | 51 (27%) | 16 (19%) | 35 (35%) | NA |

Abbreviations: BMI, body mass index; COVID-19, coronavirus disease 2019; HR, heart rate; ICU, intensive care unit; LTFU, lost to follow-up; OECD, Organisation for Economic Co-operation and Development; NA, not applicable; PCR, polymerase chain reaction; SpO2, oxygen saturation on room air; RR, respiratory rate; SARS-CoV-2, severe acute respiratory syndrome coronavirus 2.
\*Continuous variables presented as median (IQR) and compared using the Kruskal-Wallis test; categorical and binary variables presented as n(%) and compared using the Pearson $\chi^2$ test (or Fisher exact test if $n < 5$ ).
Clinical severity groups defined as: mild as having an RR $< 20$ /min and SpO<sub>2</sub> on room air $> 94\%$ at both D0 and D7; moderate disease as having a RR 20–30/minutes, SpO<sub>2</sub> 90–94% and/or receiving oxygen therapy at D0 or D7; severe disease as having a RR $> 30$ /minutes or SpO<sub>2</sub> $< 90\%$ at D0 or D7; critical disease as requiring ICU admission.
COVID-related comorbidities are based on WHO Clinical Management Guidelines and include: cardiovascular disease (including hypertension), chronic pulmonary disease (excluding asthma), renal disease, liver disease, cancer, immunosuppression (excluding HIV, including previous organ transplantation), previous psychiatric illness and dementia.
Physical measurements at D0 and D7 study visits. Oxygen saturation measured on room air if possible or retrieved from ambulance records for hospitalised participants admitted on oxygen on day of enrollment. Physical measurements not displayed for individuals with critical disease due to unreliability of measurements at admission for critically-ill patients.

We included reference samples from 37 individuals with no history of SARS-CoV-2 infection (median (IQR) age 49 years [IQR = 39.5-55.5]; 17/37 [45,9%] female). Seven (18.9%) reference samples were collected within 6 months after a primary COVID-19 vaccination series.

### COVID-19 is associated with persistent elevation of pro-inflammatory cytokines

First, we determined how the levels of cytokines in the study participants with SARS-CoV-2 infection compared to the uninfected reference group. Within the first 4 weeks after symptom onset, levels of IP10, IL10, IL17, IL1β, IL6 and TNFα were significantly elevated among participants infected with SARS-CoV-2 compared to reference samples (Supplementary Figure S1; Supplementary Table S2). Levels of IL10, IL17, IL1β and IL6 also remained elevated up to 21-24 weeks after SARS-CoV-2 infection in study participants as compared to the reference group (Supplementary Figure S1). Cytokine correlation matrices are shown in Supplementary Figure S2. These data indicate the presence of immune dysregulation in study participants with COVID-19.

### Evidence of PASC-associated immune dysregulation

We then proceeded to explore the presence of immune dysregulation among the study participants with and without PASC. At 9-12 weeks after illness onset, compared to participants without PASC, individuals with PASC tended to have lower levels of sCD14, IL10, IL17, IL1β, IL6 and TNFα in a univariable comparison (Figure 1). By 21-24 weeks, participants with PASC had significantly higher concentrations of IL10, IL1β and sCD14 than those without PASC. When restricting our analyses to participants with initially mild or moderate COVID-19, no difference in IL10 levels between participants with and without PASC remained at 21-24 weeks; however, again we observed that participants with PASC had higher levels of IL1β and sCD14 than those without PASC (Supplementary Figure S3). In univariable comparisons at 21-24 weeks, participants with an impaired diffusion capacity had significantly higher log-concentrations of IP10, IL-10, IL6 and TNFα than participants with normal diffusion capacity (Supplementary Figure S4).

**Figure 1.**
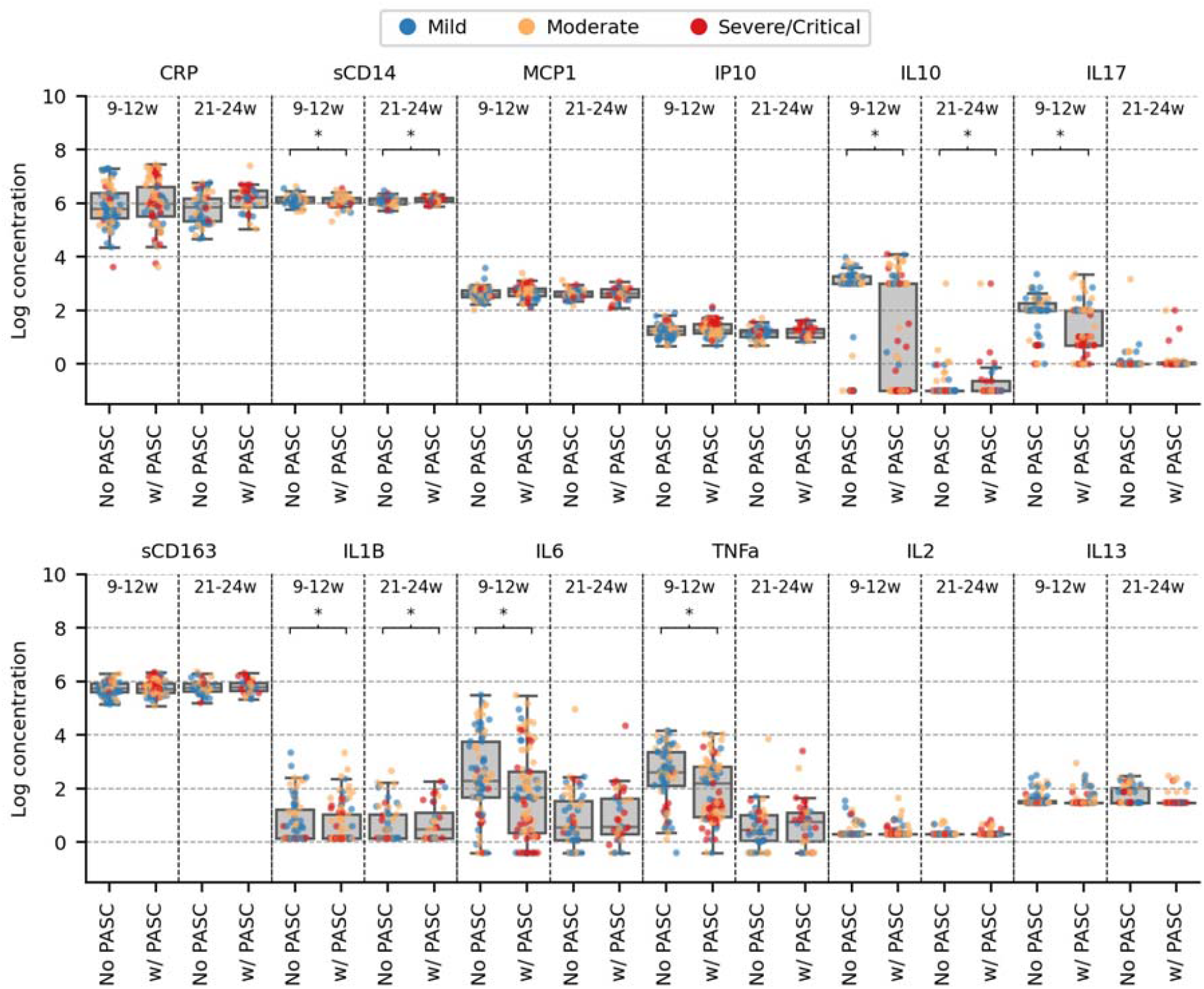
Log-concentrations of inflammatory markers at 9-12 and 21-24 weeks after Illness onset among RECoVERED study participants with and without PASC. Individuals were stratified by whether or not they reported post-acute sequelae of COVID-19 (PASC) at 12 (for measurements at 9-12 weeks) or 24 (for measurements at 21-24) weeks after Illness onset. Each dot represents an individual coloured by initial severity of COVID-19. The mean value is plotted for each individual with multiple measurements within the binned time period. A Mann-Whitney U test was used to test if measurements between those with and without PASC were significantly different. Multiple testing correction was performed using the Bonferroni method and comparisons with family-wise error rate < 0.05 were marked with an “*”.

### Factors associated with cytokine levels in multivariable models at 9-12 and 21-24 weeks after illness onset

At 9-12 weeks after disease onset, participants who had initially severe COVID-19 tended to have significantly higher levels of IP10 and sCD163 and lower levels of IL10, IL6, TNFα, IL17 and IL13 (Figure 2) compared to those with mild or moderate disease, when adjusting for other covariates. Independent of initial disease severity, having received dexamethasone during acute illness was associated with higher levels of IL6, IL10, sCD14 and CRP compared to those who did not receive dexamethasone, in multivariable analyses. Participants with PASC had significantly lower levels of IL10 and TNF-α compared to participants who had recovered from their symptoms at 9-12 weeks in multivariable analysis. Age ≥ 60 years and BMI ≥ 30 kg/m^2^ at illness onset were associated with higher levels of IL2 and IP10, and MCP1 and CRP, respectively, when adjusting for other covariates. By 21-24 weeks after illness onset, the effect of obesity and of older age on cytokine concentrations became more pronounced (Figure 3). In addition, individuals who received dexamethasone had significantly lower levels of TNFα, IL6 and IL1β by 21-24 weeks in multivariable analyses, in contrast to findings at 9-12 weeks after illness onset. Participants with ongoing PASC at 24 weeks after illness onset had higher concentrations of CRP in multivariable analyses compared to participants without PASC. When substituting PASC status for impaired diffusion capacity, we found that impaired diffusion capacity was associated with higher levels of CRP, IL6, TNFα, IP10, IL10 and IL17 in multivariable analyses (Supplementary Figure S5).

**Figure 2.**
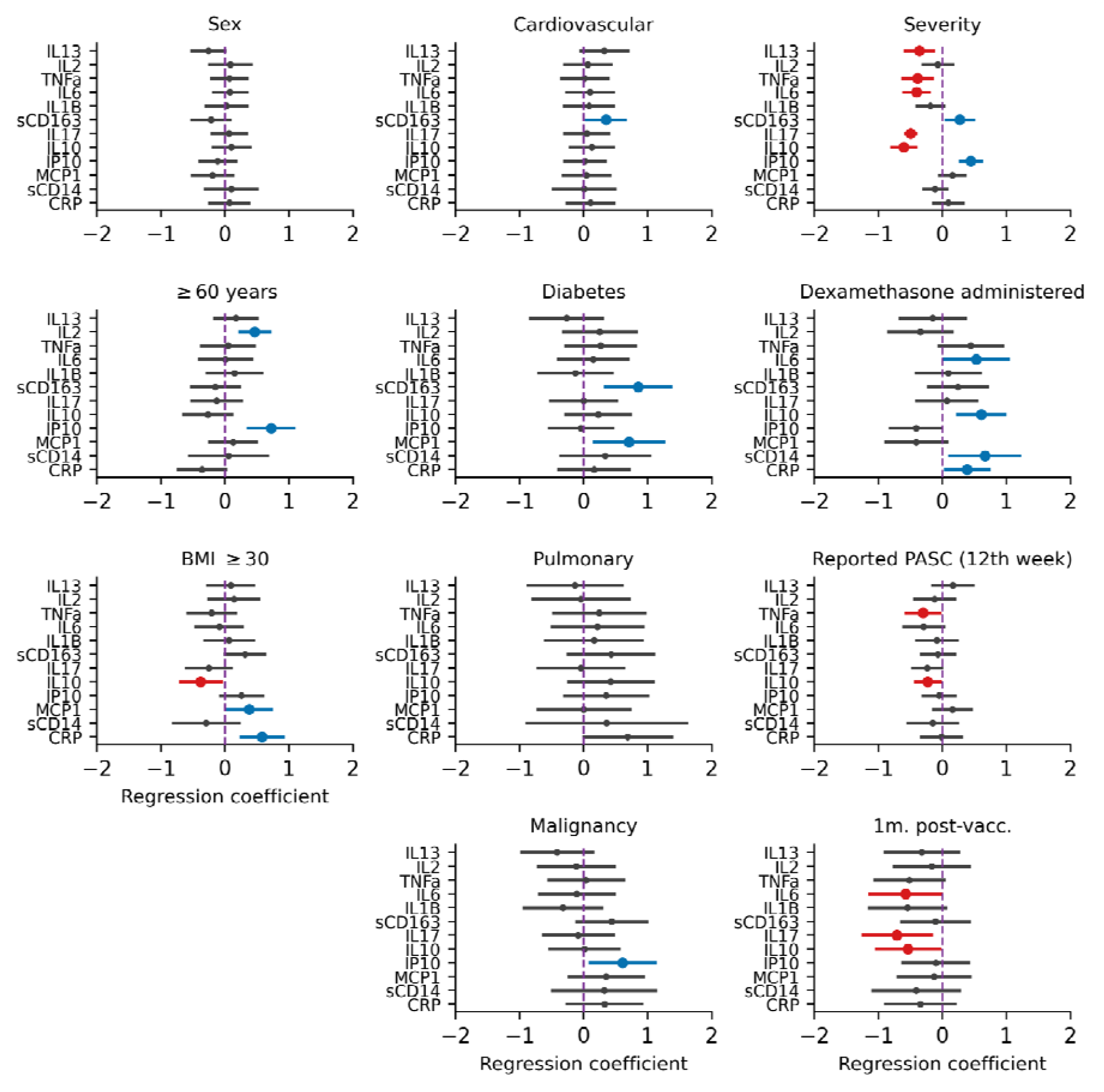
Multivariate linear regression analysis of factors associated with inflammatory marker concentrations at 9-12 weeks after COVID-19 illness onset. For each inflammatory marker, we performed a mixed effects linear regression of various factors: characteristics present prior to COVID-19 illness onset, COVID-19-related factors, and post-COVID-19 related factors. Characteristics present prior to COVID-19 illness onset included sex, age (≥60 or <60 years old), body mass index (BMI) ≥ 30, and the presence of comorbidities (including cardiovascular disease, diabetes mellitus, chronic pulmonary disease or current cancer). COVID-19-related factors included the severity of initial COVID-19 disease, if dexamethasone was administered, and current (at 12 weeks) presence of post-acute sequelae of COVID-19 (PASC). Post-COVID-19 related factors included measurement of inflammatory markers within four weeks after SARS-CoV-2 vaccination. Statistically significant negative effects (associations with lower cytokine concentrations) are shown in red whilst positive effects (associations with higher cytokine levels) are shown in blue.

**Figure 3.**
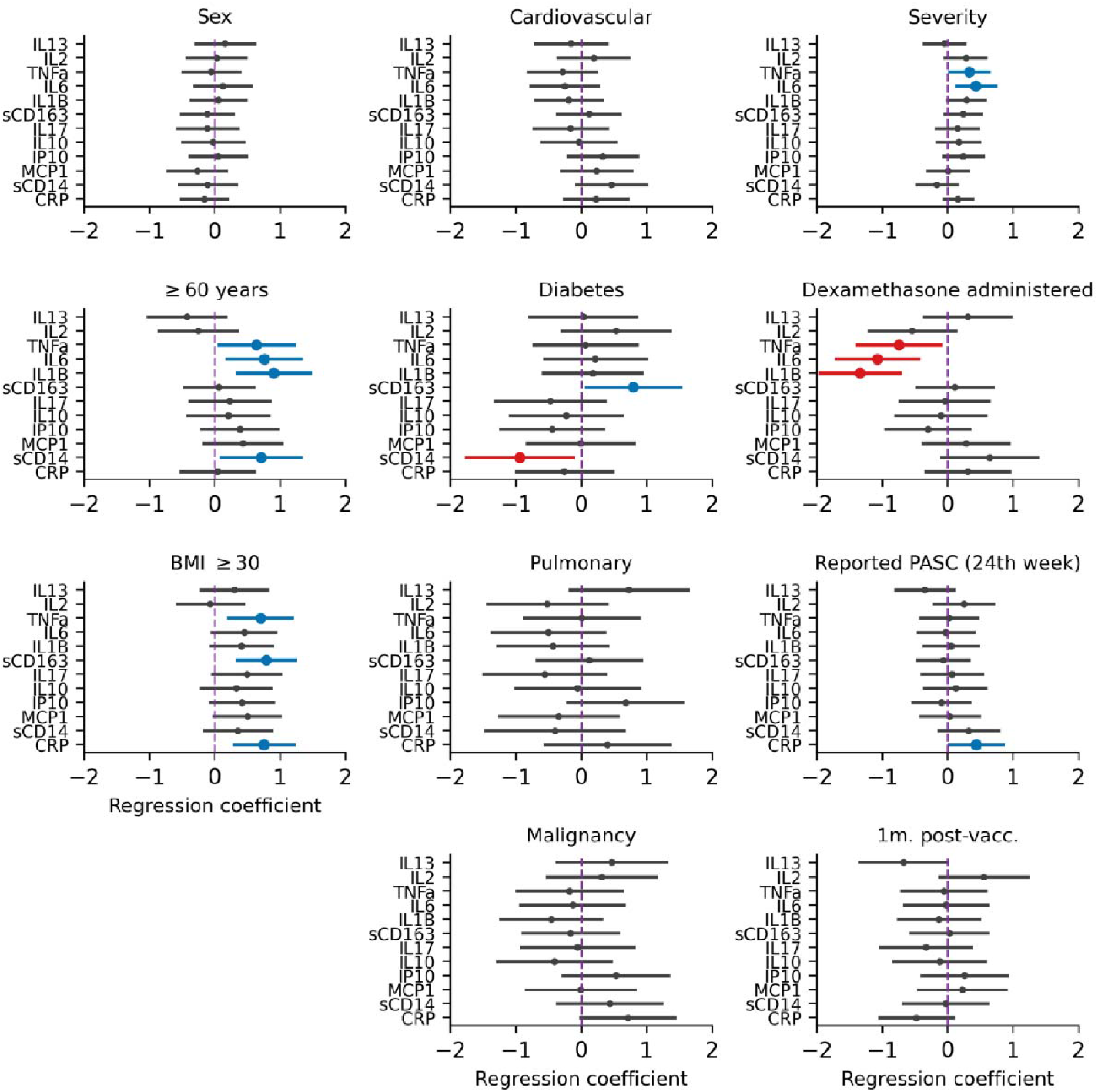
Multivariate linear regression analysis of factors associated with inflammatory marker concentrations at 21-24 weeks after COVID-19 illness onset. For each inflammatory marker, we performed a mixed effects linear regression of various factors: characteristics present prior to COVID-19 illness onset, COVID-19-related factors, and post-COVID-19 related factors. Characteristics present prior to COVID-19 illness onset included sex, age (≥60 or <60 years old), body mass index (BMI) ≥ 30, and the presence of comorbidities (including cardiovascular disease, diabetes mellitus, chronic pulmonary disease or current cancer). COVID-19-related factors included the severity of initial COVID-19 disease, if dexamethasone was administered, and current (at 24 weeks) presence of post-acute sequelae of COVID-19 (PASC). Post-COVID-19 related factors included measurement of inflammatory markers within four weeks after SARS-CoV-2 vaccination. Statistically significant negative effects (associations with lower cytokine concentrations) are shown in red whilst positive effects (associations with higher cytokine levels) are shown in blue.

### Early predictors of PASC and ongoing inflammation 6 months after illness onset

We found early IL1β and BMI at illness onset to be the strongest predictors of PASC at 21-24 weeks (Figure 4a), using Shapley additive explanation values as measures of importance[19]. Higher levels of sCD14, and to a lesser extent IL10, at 0-4 weeks were important predictors of higher levels of CRP at 21-24 weeks (Figure 4b). IL1β and TNFα measurements at 0-4 weeks were key predictors of IL6 levels at 21-24 weeks (Figure 4c). Total ANA titers were less important than BMI, initial disease severity and age in the prediction of ongoing PASC at 21-24 weeks after illness onset, but more important than sex and early levels of CRP, IL6 and IL10 (Figure 4a).

**Figure 4.**
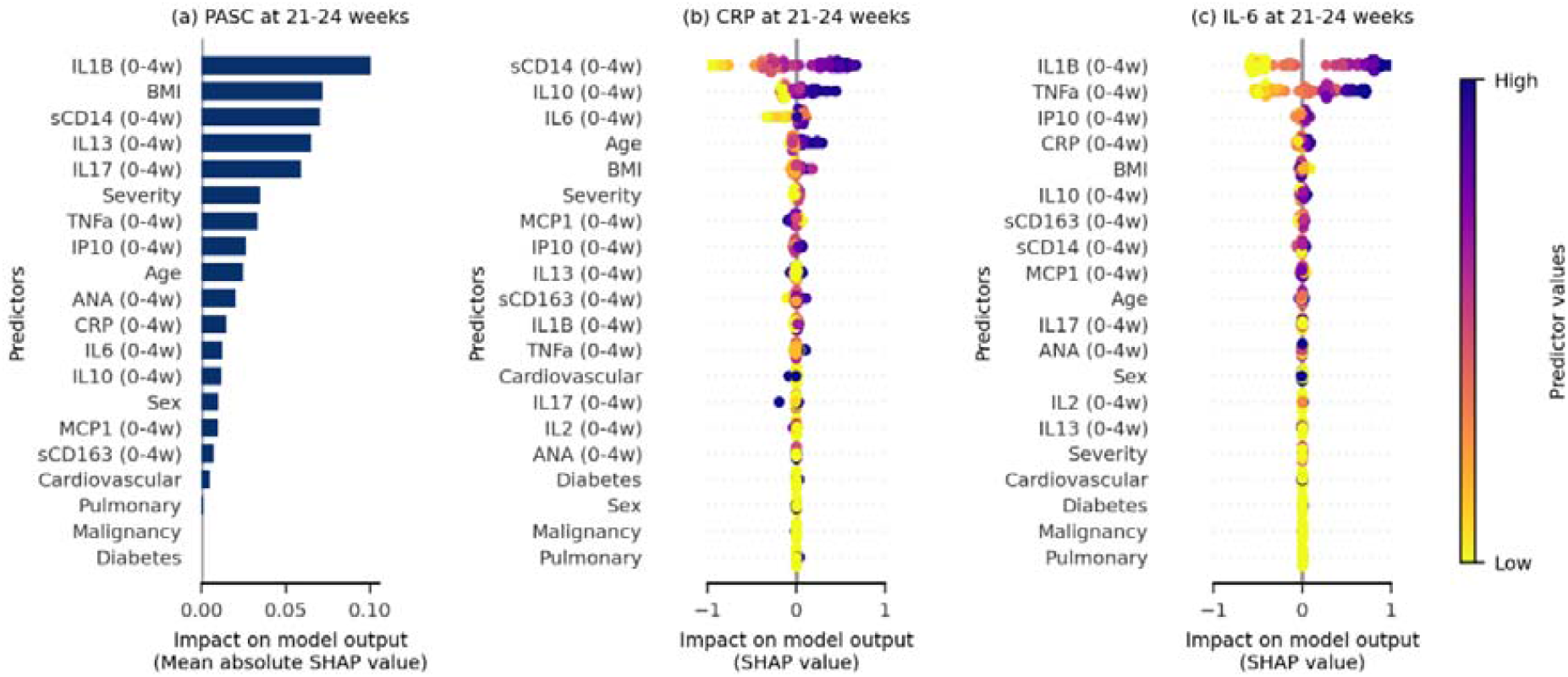
Early (0-4 week) predictors of PASC and CRP/IL-6 levels at 21-24 weeks after illness onset. Importance of different predictors (ordered from most [top] to least [bottom] important according to Shapley additive explanation (SHAP) values) on (a) individuals reporting PASC, level of (b) CRP and (c) IL-6 measurements at 21-24 weeks. Predictors include socio-demographic factors (i.e. age at infection, years, and sex), body mass index (BMI, kg/m^2^), presence/absence of comorbidities (i.e. cardiovascular disease, diabetes mellitus, chronic pulmonary disease and current cancer [malignancy]) and mean log-concentrations of inflammatory markers measured at 0-4 weeks after Illness onset.

For (a), each horizontal bar denotes the mean absolute SHAP value associated with the predictor. The larger the mean absolute SHAP value, the more important the covariate is in predicting the outcome of reporting PASC at 21-24 weeks. For (b) and (c), each point is the SHAP value for the corresponding predictor for each individual. The color of each point represents the value of the predictor for an individual. For instance, in (b), sCD14 levels at 0-4 weeks is the most important predictor for levels of CRP at 21-24 weeks, with high mean levels of sCD14 at 0-4 weeks expected to yield a higher level of CRP at 21-24 weeks.

## Discussion

In our prospective cohort study of participants with mild to critical COVID-19, we used detailed symptom data to explore cytokine profiles after disease onset and the association between PASC and cytokine profiles up to 6 months after COVID-19 illness onset. Our findings show that even months after the acute phase of disease, and regardless of PASC status, cytokine profiles are raised compared to reference samples from uninfected, healthy individuals. Although participants with PASC displayed significantly higher concentrations of pro-inflammatory CRP at 21-24 weeks after illness onset compared to those without PASC, no association was identified with other cytokines when adjusting for other factors. These observations imply that persistent inflammation cannot single-handedly explain PASC pathogenesis. However, raised IL1β at 0-4 weeks after illness onset was strongly predictive of persistent PASC at 24 weeks after illness onset, suggesting that early immune dysregulation may imprint the likelihood of developing PASC. In addition, participants with impaired diffusion capacity at 6 months after illness onset exhibited raised levels of several pro-inflammatory cytokines, also when adjusting for comorbidities and initial COVID-19 severity. This suggests that individuals with objectifiable pathology in the context of PASC may experience more pronounced immune dysregulation.

Our findings comply with many previous studies demonstrating hyperinflammation during acute COVID-19[21-24], and indicate that elevated levels of proinflammatory cytokines may persist for several months after initial infection. However, our observations among participants with PASC were inconclusive. We did not find clear evidence that PASC, as defined by self-reported symptoms, is driven by prolonged inflammation (for instance, due to viral persistence), as has been speculated[25] and demonstrated in several previous studies that illustrated increased levels of pro-inflammatory cytokines such as IL6, IL1β and TNFα among individuals with PASC [6, 13, 14]. In our analysis, when adjusting for age, sex, comorbidities (including obesity) at illness onset, initial COVID-19 severity, dexamethasone treatment and recent COVID-19 vaccination, PASC was associated only with raised CRP at 21-24 weeks after illness onset. However, when substituting PASC status (based on self-reported symptoms) with impaired diffusion capacity as an objective measure, the pattern of raised pro-inflammatory cytokines (including IL10, IL6, IL17, IP10 and TNFα) observed in other studies was reiterated. This observation may reflect two possible explanations. Firstly, that confirming PASC status with an objective measure increased the precision of the definition, reducing misclassification bias resulting from self-reported symptoms not related to PASC and thus allowing an association with pro-inflammatory cytokines to be revealed. Alternatively, PASC is an umbrella term for multiple conditions, within which individuals with measurable lung abnormalities experience a persistent hyperinflammatory process whilst the underlying cause of those with other symptom clusters[27] cannot be explained by aberrant cytokines. Our findings thus suggest that immune dysregulation plays an important role in PASC pathogenesis in some individuals, and that those with measurable persistent pathology following COVID-19 exhibit prolonged hyperinflammation.

We also observed that higher levels of IL1β (and to a lesser extent sCD14, IL13, IL17 and TNFα) at 0-4 weeks were strongly associated with ongoing PASC at 24 weeks after illness onset. This is of interest because we did not find an association of PASC with these cytokines at 21-24 weeks, suggesting that early IL1β induced tissue damage[30] or endothelial dysfunction[31] may predispose individuals to later PASC. In addition, we explored early predictors for the proinflammatory cytokine levels (CRP and IL-6) at 24 weeks, to overcome any error in our clinical measurement of PASC. Elevated early sCD14 and IL10 levels were most strongly predictive of raised CRP at 21-24 weeks, whilst IL1β and TNFα levels were strongly congruent with persistently elevated IL6 at 24 weeks. These findings help provide insight into the possible immunopathogenesis of PASC. First, the association between high sCD14 levels at 0-4 weeks and persistently elevated CRP and PASC at 24 weeks suggests the key role of monocyte-macrophage activation in the acute phase[32], driving long-term inflammation. Second, early IL1β was associated with both PASC and raised IL-6 at 6 months, implying that IL1β-mediated acute inflammation contributes to ongoing symptomatology and hyperinflammation many months after infection. Given that IL1β has been implicated as a marker of neuroinflammation[33] and profound neurological symptoms are part of PASC[34], further exploration of the role of this cytokine in PASC pathogenesis is warranted. Consistent with previous findings[15], higher BMI at illness onset was also an independent predictor of ongoing PASC at 24 weeks. This suggests that the effect of BMI on PASC risk is not only mediated by inflammation (as indicated by the association of obesity at illness onset with raised TNFα, sCD163 and CRP at 21-24 weeks), but also by other physiological and metabolic processes[35]. It is crucial that ongoing studies on the role of inflammation in PASC pathogenesis account for the central role played by BMI.

Our study benefits from its prospective design, thus limiting selection bias, detailed symptom data, representation of a wide range of COVID-19 severity, and the use of reference values from SARS-CoV-2-naïve individuals without comorbidities. However, our study also has limitations. Firstly, we did not have symptom data pre-dating SARS-CoV-2 infection. We therefore cannot be certain that reported symptoms were a result of COVID-19 or due to pre-existing comorbidities, which may have resulted in misclassification bias. However, we attempted to overcome this bias by defining long COVID symptoms as those with a date of onset within 1 month from overall COVID-19 onset. In addition, using lung function results as an objective measure of PASC with respiratory sequelae PASC allowed us to reduce noise from self-reported symptoms not driven by PASC pathology. Another limitation was that our reference group consisted of individuals without comorbidities. As such, we are unable to ascertain to what extent differences observed in cytokine levels in cohort participants were associated with underlying comorbidities. This limitation is compounded by the fact that there were no pre-COVID serum samples available to distinguish any pre-existing aberrant cytokine levels resulting from underlying comorbidities. Finally, although electronic patient records were used to verify comorbidities for hospitalised participants, we did not have access to primary care records for non-hospitalised participants. It was reassuring to observe raised levels of sCD163 over time in participants with diabetes, an association that has previously been reported[36], suggesting response bias for self-reported comorbidities was limited.

## Conclusions

In summary, our study indicates that immune dysregulation does not single-handedly explain PASC as defined by self-reported symptoms. However, confirmation of PASC status with impaired pulmonary function as an objective measure revealed an association with raised pro-inflammatory cytokines, as previously reported in other studies. In addition, early raised IL1β levels were strongly predictive of ongoing PASC at 6 months in our analyses. Our findings therefore suggest that immune dysregulation plays an important role in the pathogenesis of ongoing symptoms in some individuals.

## Data Availability

All data produced in the present study are available upon reasonable request to the authors

## Acknowledgments

**RECoVERED Study Group:** *<u>From the Public Health Service of Amsterdam</u>*: Ivette Agard, Jane Ayal, Floor Cavdar, Marianne Craanen, Udi Davinovich, Annemarieke Deuring, Annelies van Dijk, Maartje Dijkstra, Ertan Ersan, Laura del Grande, Joost Hartman, Nelleke Koedoot, Romy Lebbink, Tjalling Leenstra, Dominique Loomans, Agata Makowska, Tom du Maine, Ilja de Man, Amy Matser, Lizenka van der Meij, Marleen van Polanen, Maria Oud, Clark Reid, Leeann Storey, Marije de Wit, Marc van Wijk. *<u>From Amsterdam University Medical Centers</u>*: Joyce van Assem, Marijne van Beek, Orlane Figaroa, Leah Frenkel, Agnes Harskamp-Holwerda, Mette Hazenberg, Soemeja Hidad, Nina de Jong, Hans Knoop, Lara Kuijt, Anja Lok, Eric Moll van Charante, Colin Russell, Annelou van der Veen, Bas Verkaik, Gerben-Rienk Visser.

The authors wish to thank all RECoVERED study participants.

