## Supplementary Materials for "Inflammatory profiles are associated with long COVID up to 6 months after illness onset: a prospective cohort study of individuals with mild to critical COVID-19"

### Tables

**Supplementary Table S1.** Socio-demographic, clinical and COVID-19-related characteristics of RECoVERED participants included and excluded in the current analyses

**Supplementary Table S2.** Concentration distribution of cytokines across different time-points among individuals with COVID-19 and healthy controls

### Figures

**Supplementary Figure S1.** Log-concentrations of inflammatory markers over time since Illness onset among RECoVERED study participants, compared to healthy uninfected controls

**Supplementary Figure S2.** Correlation matrices of inflammatory markers at 0-4, 9-12 and 21-24 weeks after illness onset

**Supplementary Figure S3.** Log-concentrations of inflammatory markers at 9-12 and 21-24 weeks after Illness onset among RECoVERED study participants with initially mild or moderate COVID-19 only, stratified by PASC status

**Supplementary Figure S4.** Log-concentrations of inflammatory markers at 21-24 weeks after Illness onset among RECoVERED study participants with, stratified by impaired diffusion capacity at month 6 after illness onset

**Supplementary Figure S5.** Multivariate linear regression analysis of factors associated with inflammatory marker concentrations at 9-12 weeks after COVID-19 illness onset

**Supplementary Table S1. Socio-demographic, clinical and COVID-19-related characteristics of RECoVERED participants included and excluded in the current analyses**

|  | **Total** | **Excluded** | **Included** | **p-value** |
| --- | --- | --- | --- | --- |
|  | **N=349** | **N=163** | **N=186** |  |
| Sex |  |  |  | 0.007 |
| Male | 196 (56%) | 79 (48%) | 117 (63%) |  |
| Female | 153 (44%) | 84 (52%) | 69 (37%) |  |
| Age, years | 51.0 (36.0-62.0) | 49.0 (33.0-61.0) | 52.0 (37.0-62.0) | 0.31 |
| BMI, kg/m^2^ | 26.2 (23.4-29.7) | 27.1 (23.5-30.7) | 25.7 (23.2-29.3) | 0.25 |
| BMI category |  |  |  | 0.092 |
| Normal weight | 141 (40%) | 66 (40%) | 75 (40%) |  |
| Overweight | 114 (33%) | 43 (26%) | 71 (38%) |  |
| Obese | 83 (24%) | 44 (27%) | 39 (21%) |  |
| Missing | 11 (3%) | 10 (6%) | 1 (1%) |  |
| Migration background |  |  |  | 0.033 |
| Dutch | 193 (55%) | 69 (42%) | 124 (67%) |  |
| Non-Dutch, OECD high-income | 40 (11%) | 18 (11%) | 22 (12%) |  |
| Non-Dutch, OECD low/middle income | 80 (23%) | 42 (26%) | 38 (20%) |  |
| Missing | 36 (10%) | 34 (21%) | 2 (1%) |  |
| Smoking |  |  |  | 0.47 |
| Non-smoker | 206 (59%) | 96 (59%) | 110 (59%) |  |
| Smoker | 22 (6%) | 10 (6%) | 12 (6%) |  |
| Ex-smoker | 102 (29%) | 40 (25%) | 62 (33%) |  |
| Missing | 19 (5%) | 17 (10%) | 2 (1%) |  |
| Highest level of education |  |  |  | 0.011 |
| None, primary or secondary education | 45 (13%) | 18 (11%) | 27 (15%) |  |
| Vocational training | 77 (22%) | 42 (26%) | 35 (19%) |  |
| University education | 185 (53%) | 64 (39%) | 121 (65%) |  |
| Missing | 42 (12%) | 39 (24%) | 3 (2%) |  |
| Number of COVID-19 high-risk comorbidities |  |  |  | 0.035 |
| 0 | 189 (54%) | 87 (53%) | 102 (55%) |  |
| 1 | 81 (23%) | 30 (18%) | 51 (27%) |  |
| 2 | 49 (14%) | 31 (19%) | 18 (10%) |  |
| 3 or more | 30 (9%) | 15 (9%) | 15 (8%) |  |
| Cardiovascular disease | 95 (28%) | 48 (31%) | 47 (25%) | 0.27 |
| Diabetes | 46 (13%) | 30 (19%) | 16 (9%) | 0.004 |
| Chronic respiratory disease | 26 (8%) | 14 (9%) | 12 (6%) | 0.39 |
| Cancer | 18 (5%) | 6 (4%) | 12 (6%) | 0.28 |
| Immunosuppressed | 6 (2%) | 2 (1%) | 4 (2%) | 0.54 |
| Psychiatric illness | 19 (6%) | 7 (4%) | 12 (6%) | 0.42 |
| Other comorbidities | 78 (23%) | 37 (24%) | 41 (22%) | 0.67 |

|  | **Total** | **Excluded** | **Included** | **p-value** |
| --- | --- | --- | --- | --- |
|  | **N=349** | **N=163** | **N=186** |  |
| Clinical severity score |  |  |  | 0.011 |
| Mild | 99 (28%) | 35 (21%) | 64 (34%) |  |
| Moderate | 151 (43%) | 72 (44%) | 79 (42%) |  |
| Severe/critical | 99 (28%) | 56 (34%) | 43 (23%) |  |
| Hospital admission | 179 (51%) | 105 (64%) | 74 (40%) | <0.001 |
| ICU admission | 45 (13%) | 19 (12%) | 26 (14%) | 0.63 |
| Days from illness onset to COVID-19 diagnosis | 4 (2-10) | 4 (2-11) | 5 (2-9) | 0.43 |
| Days from illness onset to hospitalisation | 9 (7-14) | 9 (6-14) | 9 (7-12) | 0.90 |
| Days from illness onset to ICU admission | 10 (7-12) | 9 (7-12) | 10 (7-11) | 0.71 |
| Received oxygen therapy before or during follow-up | 169 (49%) | 97 (61%) | 72 (39%) | <0.001 |
| Type of steroid |  |  |  | <0.001 |
| No steroid | 241 (69%) | 98 (60%) | 143 (77%) |  |
| Dexamethasone | 79 (23%) | 54 (33%) | 25 (13%) |  |
| Other steroid | 28 (8%) | 10 (6%) | 18 (10%) |  |
| Missing | 1 (0%) | 1 (1%) | 0 (0%) |  |
| Maximal HR, beats/min | 83 (72-94) | 87 (76-97) | 79 (71-92) | 0.002 |
| Maximal RR, breaths/min | 20 (16-24) | 20 (16-24) | 20 (16-22) | 0.050 |
| Lowest SpO_2_, % | 96 (91-98) | 95 (89-98) | 96 (92-98) | 0.040 |
| COVID-19 vaccination status (primary series) |  |  |  | <0.001 |
| Not vaccinated | 31 (9%) | 30 (18%) | 1 (1%) |  |
| Vaccinated | 228 (65%) | 43 (26%) | 185 (99%) |  |
| LTFU before vaccination | 90 (26%) | 90 (55%) | 0 (0%) |  |
| Time from illness onset to first vaccination, days | 244 (144-361) | 247 (77-345) | 244 (151-363) | 0.46 |
| Died during follow-up | 5 (1%) | 4 (2%) | 1 (1%) | 0.19 |
| Place of recruitment |  |  |  | <0.001 |
| Non-hospital | 161 (46%) | 55 (34%) | 106 (57%) |  |
| Hospital | 188 (54%) | 108 (66%) | 80 (43%) |  |
| Type of inclusion |  |  |  | 0.81 |
| Prospective | 257 (74%) | 121 (74%) | 136 (73%) |  |
| Retrospective | 92 (26%) | 42 (26%) | 50 (27%) |  |
| Days from illness onset to inclusion in study | 12 (6-38) | 14 (8-32) | 10 (5-62) | 0.14 |
| Prospective inclusions only | 9 (5-15) | 11 (7-16) | 8 (4-13) | <0.001 |
| Retrospective inclusions only | 85 (72-94) | 79 (52-94) | 87 (81-94) | 0.086 |
| Follow-up time from enrolment in study | 499.0 (274.0-659.0) | 274.0 (85.0-499.0) | 581.0 (486.0-696.0) | <0.001 |
| Lost to follow-up | 158 (45%) | 107 (66%) | 51 (27%) | NA |

Abbreviations: BMI, body mass index; COVID-19, coronavirus disease 2019; HR, heart rate; ICU, intensive care unit; LTFU, lost to follow-up; OECD, Organisation for Economic Co-operation and Development; NA, not applicable; PCR, polymerase chain reaction; SpO2, oxygen saturation on room air; RR, respiratory rate; SARS-CoV-2, severe acute respiratory syndrome coronavirus 2.

*Continuous variables presented as median (IQR) and compared using the Kruskal-Wallis test; categorical and binary variables presented as n(%) and compared using the Pearson χ 2 test (or Fisher exact test if n <5).

Clinical severity groups defined as: mild as having an RR <20/min and SpO2 on room air >94% at both D0 and D7; moderate disease as having a RR 20–30/minutes, SpO2 90–94% and/or receiving oxygen therapy at D0 or D7; severe disease as having a RR >30/minutes or SpO2 <90% at D0 or D7; critical disease as requiring ICU admission.

COVID-related comorbidities are based on WHO Clinical Management Guidelines and include: cardiovascular disease (including hypertension), chronic pulmonary disease (excluding asthma), renal disease, liver disease, cancer, immunosuppression (excluding HIV, including previous organ transplantation), previous psychiatric illness and dementia.

Physical measurements (HR, RR and SpO_2_) at D0 and D7 study visits. Oxygen saturation measured on room air if possible or retrieved from ambulance records for hospitalized participants admitted on oxygen on day of enrollment.

**Supplementary Table S2. Concentration distribution of soluble inflammatory markers across different time-points among individuals with COVID-19 and health controls**

|  |  | **Persons with COVID-19** | | |  | **Healthy controls** | | |
| --- | --- | --- | --- | --- | --- | --- | --- | --- |
| **Marker** | **Weeks since symptom onset** | **Median** | **25th percentile** | **75th percentile** | ***p**** | **Median** | **25th percentile** | **75th percentile** |
| CRP  (log pg/ml) | ≤4 | 6.39 | 5.86 | 7.21 | 0.094 | 6.07 | 5.80 | 6.36 |
|  | 9-12 | 5.93 | 5.46 | 6.45 | 0.432 |  |  |  |
|  | 21-24 | 5.99 | 5.51 | 6.37 | 0.432 |  |  |  |
| sCD14  (log pg/ml) | ≤4 | 6.27 | 6.12 | 6.38 | p<0.001 | 6.72 | 6.34 | 6.95 |
|  | 9-12 | 6.13 | 6.01 | 6.21 | p<0.001 |  |  |  |
|  | 21-24 | 6.09 | 6.00 | 6.18 | p<0.001 |  |  |  |
| MCP1  (log pg/ml) | ≤4 | 2.73 | 2.59 | 2.87 | 0.351 | 2.66 | 2.60 | 2.75 |
|  | 9-12 | 2.65 | 2.54 | 2.80 | 0.48 |  |  |  |
|  | 21-24 | 2.60 | 2.51 | 2.77 | 0.202 |  |  |  |
| IP-10  (log pg/ml) | ≤4 | 1.83 | 1.60 | 2.05 | p<0.001 | 1.23 | 1.12 | 1.32 |
|  | 9-12 | 1.27 | 1.11 | 1.45 | 0.48 |  |  |  |
|  | 21-24 | 1.17 | 1.00 | 1.28 | 0.094 |  |  |  |
| IL-10  (log pg/ml) | ≤4 | 3.86 | 3.10 | 4.14 | p<0.001 | -1.00 | -1.00 | -1.00 |
|  | 9-12 | 3.00 | -1.00 | 3.10 | p<0.001 |  |  |  |
|  | 21-24 | -1.00 | -1.00 | -0.79 | 0.006 |  |  |  |
| IL-17  (log pg/ml) | ≤4 | 2.00 | 2.00 | 2.45 | p<0.001 | 0.00 | 0.00 | 0.00 |
|  | 9-12 | 2.00 | 0.70 | 2.00 | p<0.001 |  |  |  |
|  | 21-24 | 0.00 | 0.00 | 0.00 | 0.04 |  |  |  |
| sCD163  (log pg/ml) | ≤4 | 5.79 | 5.60 | 5.98 | 0.202 | 5.65 | 5.53 | 5.88 |
|  | 9-12 | 5.73 | 5.60 | 5.93 | 0.432 |  |  |  |
|  | 21-24 | 5.78 | 5.64 | 5.93 | 0.265 |  |  |  |
| IL-1𝛃  (log pg/ml) | ≤4 | 0.85 | 0.15 | 1.70 | p<0.001 | 0.15 | 0.15 | 0.15 |
|  | 9-12 | 0.15 | 0.15 | 1.11 | p<0.001 |  |  |  |
|  | 21-24 | 0.15 | 0.15 | 1.04 | p<0.001 |  |  |  |
| IL-6  (log pg/ml) | ≤4 | 3.38 | 2.55 | 4.32 | p<0.001 | 0.50 | 0.21 | 0.50 |
|  | 9-12 | 2.00 | 0.86 | 3.12 | p<0.001 |  |  |  |
|  | 21-24 | 0.59 | 0.08 | 1.60 | 0.047 |  |  |  |
| TNF-𝛂  (log pg/ml) | ≤4 | 3.21 | 2.88 | 3.69 | p<0.001 | 0.36 | 0.36 | 0.73 |
|  | 9-12 | 2.48 | 1.13 | 2.93 | p<0.001 |  |  |  |
|  | 21-24 | 0.57 | 0.06 | 1.03 | 0.283 |  |  |  |
| IL-2  (log pg/ml) | ≤4 | 0.30 | 0.30 | 0.54 | 0.202 | 0.30 | 0.30 | 0.30 |
|  | 9-12 | 0.30 | 0.30 | 0.30 | 0.005 |  |  |  |
|  | 21-24 | 0.30 | 0.30 | 0.30 | p<0.001 |  |  |  |
| IL-13  (log pg/ml) | ≤4 | 1.48 | 1.48 | 1.91 | 0.477 | 1.48 | 1.48 | 1.48 |
|  | 9-12 | 1.48 | 1.48 | 1.48 | 0.265 |  |  |  |
|  | 21-24 | 1.48 | 1.48 | 1.88 | 0.004 |  |  |  |

*Mann-Whitney U test between measurements at each binned time period of infected individuals and those measured from healthy controls with multiple testing correction using the Bonferroni method.

**Supplementary Figure S1**. **Log-concentrations of inflammatory markers over time since Illness onset among RECoVERED study participants, compared to healthy uninfected controls**

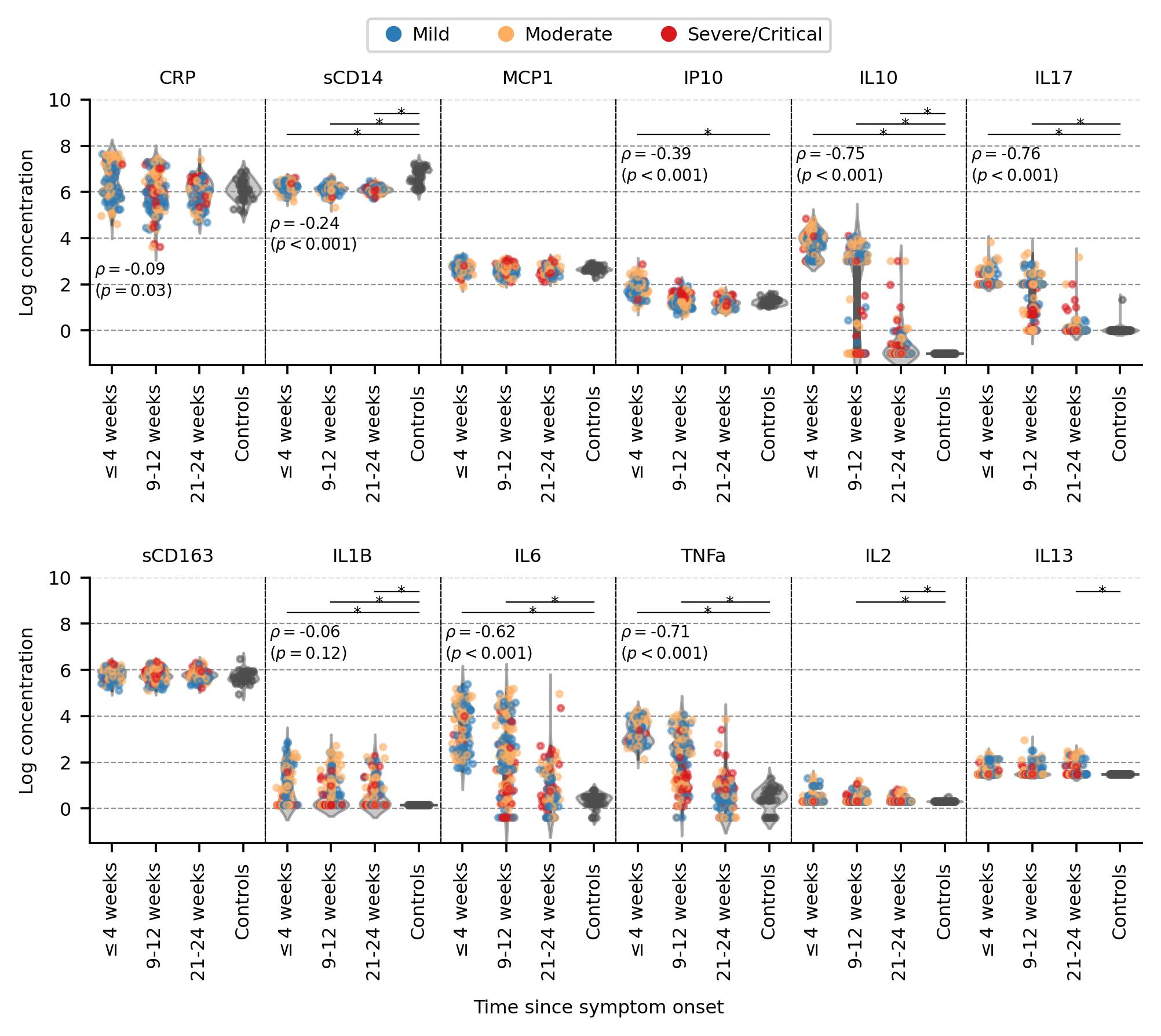

Each dot represents an individual coloured by the severity of their COVID-19 infection. Time is binned into: within the first four weeks since symptom onset, 9-12 weeks, and 12-24 weeks after symptom onset. The mean measurement value is plotted for each individual with multiple measurements within the binned time period. Measurements from 37 healthy controls who were not infected with SARS-CoV-2 (in gray) were also plotted. The Mann-Whitney U test was used to test if measurements at each binned time period of infected individuals are significantly different from those measured from healthy controls. Multiple testing correction was performed using the Bonferroni method and comparisons with family-wise error rate < 0.05 were marked with an “*”. If concentration of inflammatory markers of infected individuals during the first four weeks after symptom onset are significantly different from those measured from controls, we additionally computed the Spearman rank correlation coefficient ($\rho$) and its corresponding p-value to assess its monotonic relationship against time.

**Supplementary Figure S2. Correlation matrices of inflammatory markers at 0-4, 9-12 and 21-24 weeks after illness onset**

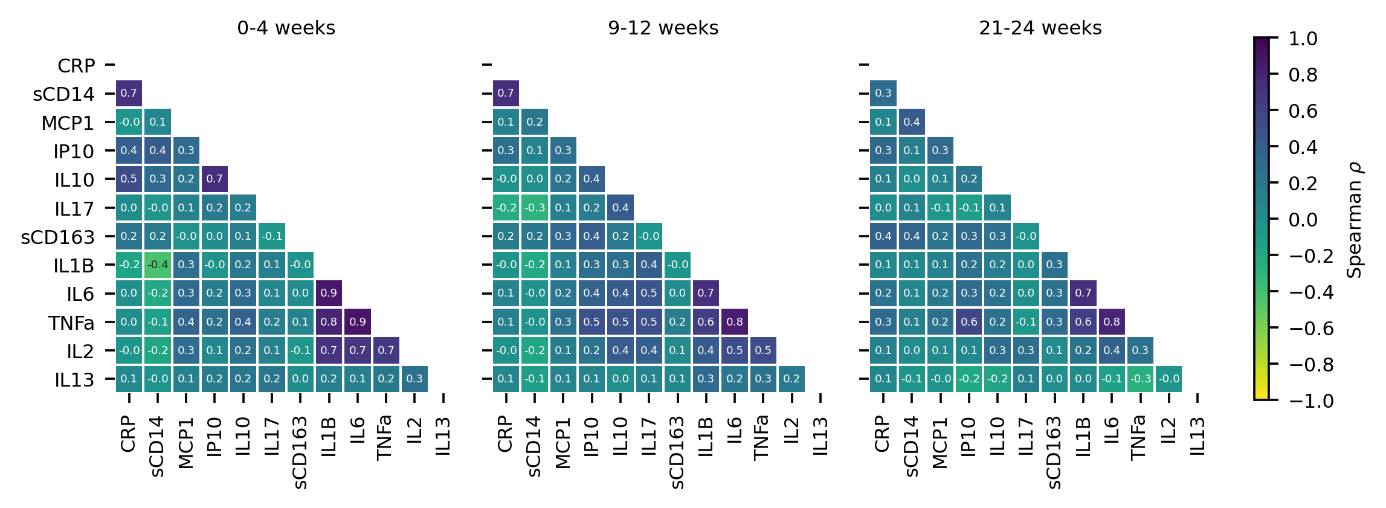

**Supplementary Figure S3. Log-concentrations of inflammatory markers at 9-12 and 21-24 weeks after Illness onset among RECoVERED study participants with initially mild or moderate COVID-19 only, stratified by PASC status**

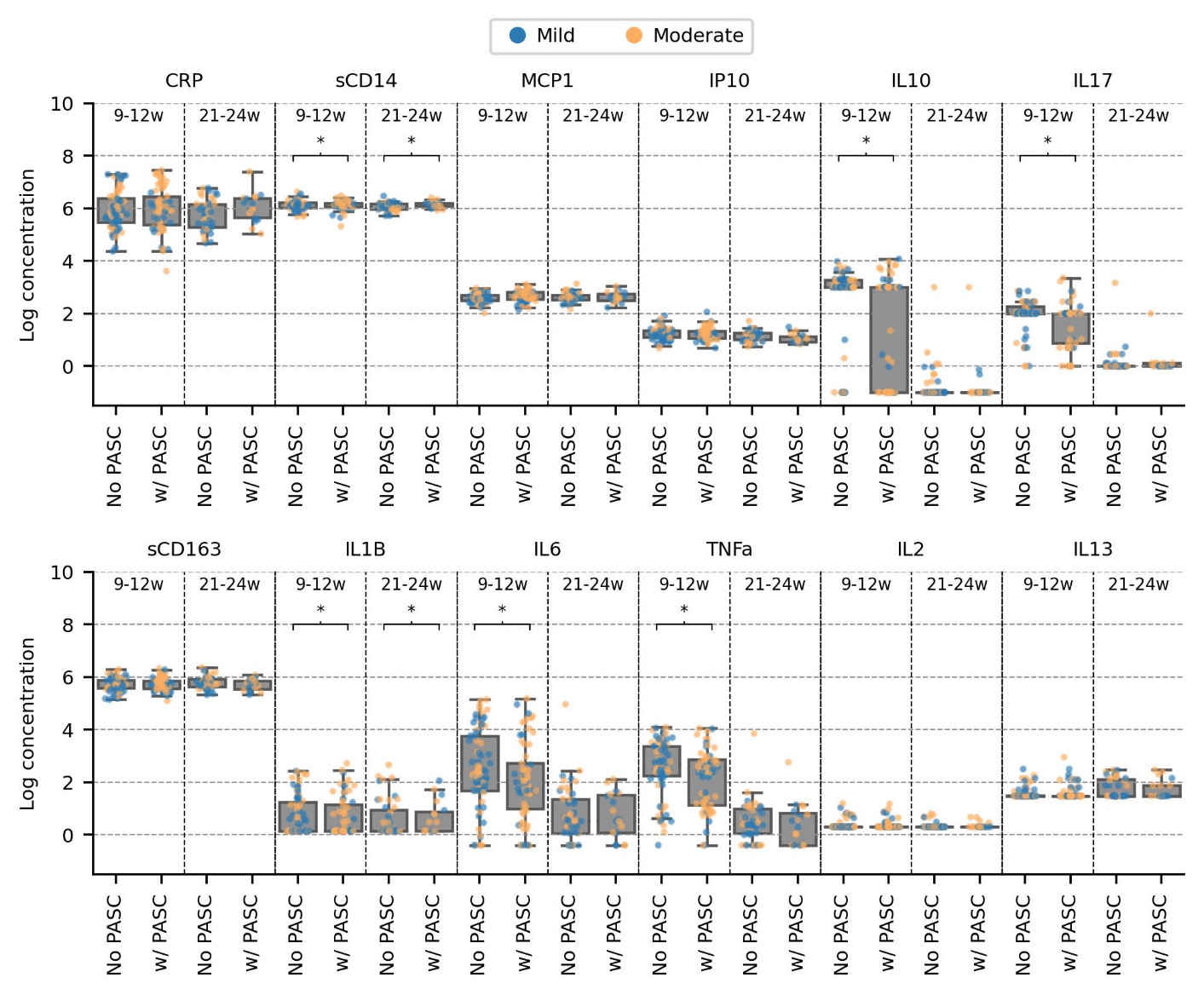

Individuals were stratified by whether or not they reported post-acute sequelae of COVID-19 (PASC) at 12 (for measurements at 9-12 weeks) or 24 (for measurements at 21-24) weeks after Illness onset. Each dot represents an individual coloured by initial severity of COVID-19 (mild or moderate disease only). The mean value is plotted for each individual with multiple measurements within the binned time period. A Mann-Whitney U test was used to test if measurements between those with and without PASC were significantly different. Multiple testing correction was performed using the Bonferroni method and comparisons with family-wise error rate < 0.05 were marked with an “*”.

**Supplementary Figure S4. Log-concentrations of inflammatory markers at 21-24 weeks after illness onset among RECoVERED study participants, stratified by impaired diffusion capacity at month 6 after illness onset**

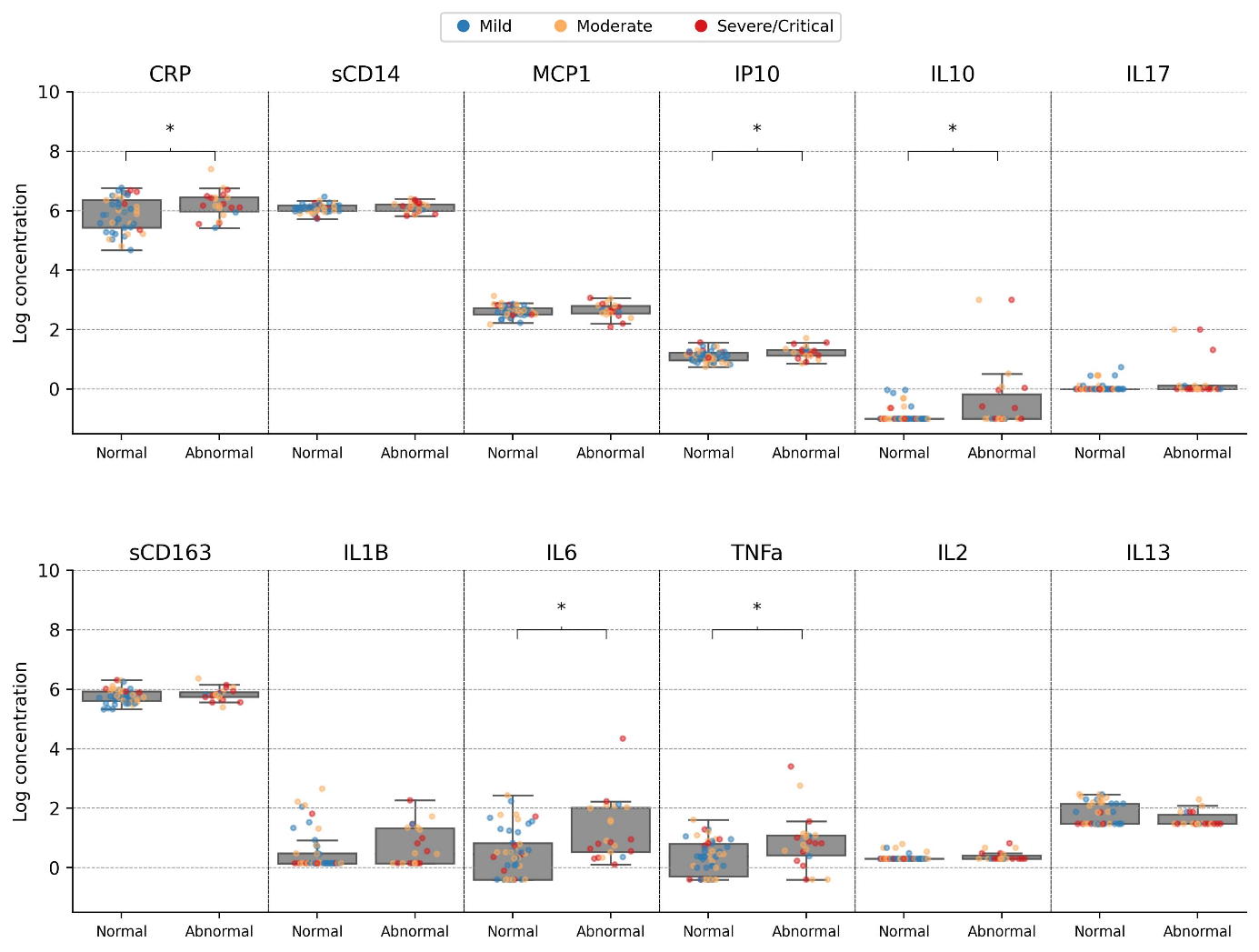

Individuals were stratified by whether or not their lung function test was normal or abnormal (abnormal defined as having an impaired diffusion capacity [D_LCO_] – i.e., below the lower limit of normal). Each dot represents an individual coloured by initial severity of COVID-19 (mild or moderate disease only). The mean value is plotted for each individual with multiple measurements within the binned time period. A Mann-Whitney U test was used to test if measurements between those with and without abnormal lung function were significantly different. Multiple testing correction was performed using the Bonferroni method and comparisons with family-wise error rate < 0.05 were marked with an “*”.

**Supplementary Figure S5. Multivariate linear regression analysis of factors associated with inflammatory marker concentrations at 21-24 weeks after COVID-19 illness onset, including abnormal lung function (impaired diffusion capacity)**

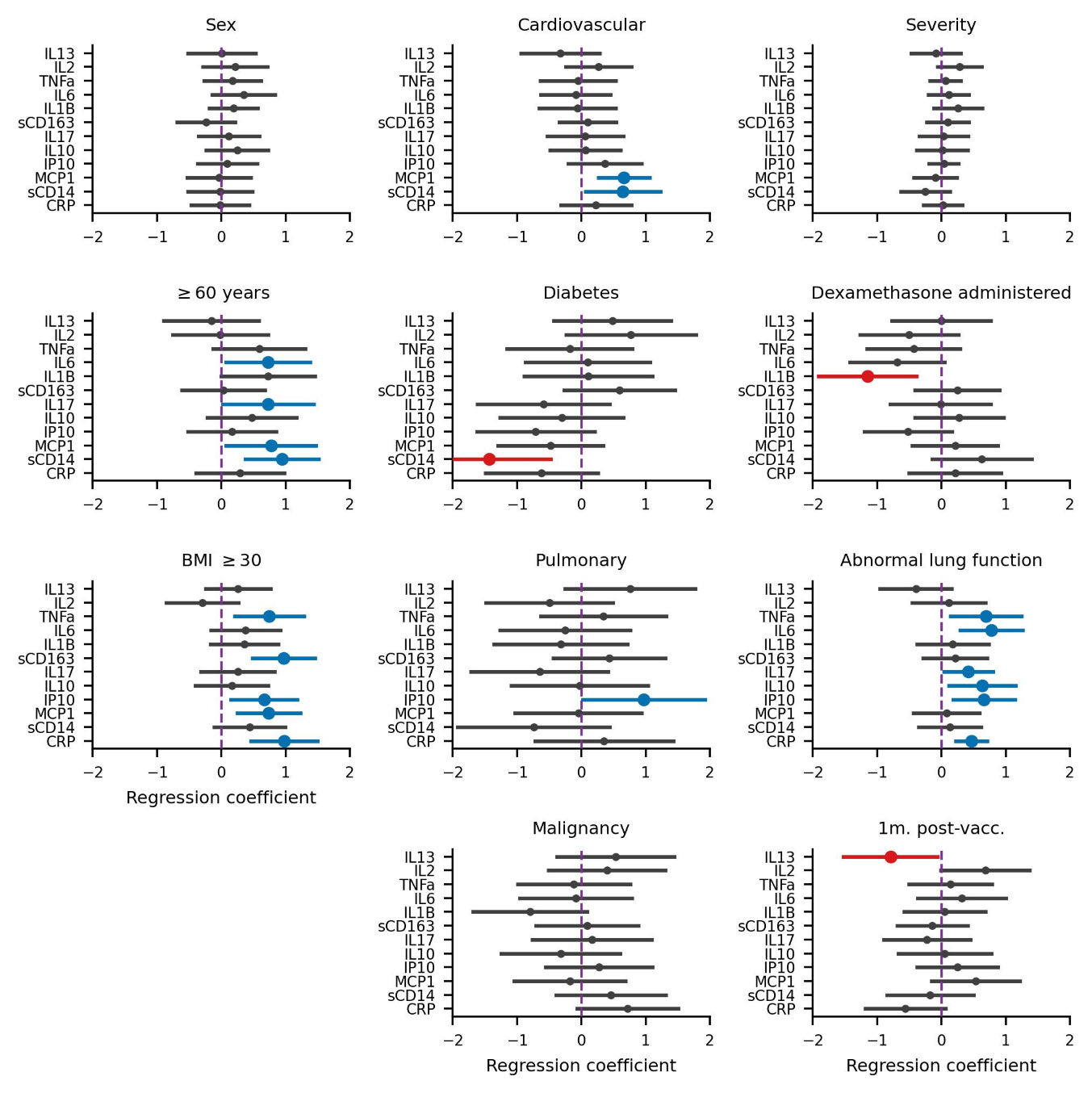

For each inflammatory marker, we performed a mixed effects linear regression of various factors: characteristics present prior to COVID-19 illness onset, COVID-19-related factors, and post-COVID-19 related factors. Characteristics present prior to COVID-19 illness onset included sex, age (≥60 or <60 years old), body mass index (BMI) ≥ 30, and the presence of comorbidities (including cardiovascular disease, diabetes mellitus, chronic pulmonary disease or current cancer). COVID-19-related factors included the severity of initial COVID-19 disease, if dexamethasone was administered, and current (at 24 weeks) presence of abnormal lung function (impaired diffusion capacity [D_LCO_]). Post-COVID-19 related factors included measurement of inflammatory markers within four weeks after SARS-CoV-2 vaccination. Statistically significant negative effects (associations with lower cytokine concentrations) are shown in red whilst positive effects (associations with higher cytokine levels) are shown in blue.
